# Intermittent fasting alters tumor burden, autophagy, and metabolites in chronic lymphocytic leukemia

**DOI:** 10.1101/2024.12.16.24319108

**Authors:** Eleah Stringer, Zhengxiao Wei, Samantha Punch, Nathalie Costie, Jun Han, David Goodlett, Farouk S. Nathoo, Julian J. Lum, Nicol Macpherson

**Affiliations:** Department of Oncology Nutrition, BC Cancer- Victoria, Victoria, BC, Canada, V8R 6V5; Department of Health Information Science, University of Victoria, Victoria, BC, Canada, V8W 2Y2; Department of Mathematics and Statistics, University of Victoria, Victoria, BC, Canada, V8W 2Y2; Trev and Joyce Deeley Research Centre, BC Cancer, Victoria, BC, Canada, V8R 6V5; University of Victoria Genome BC Proteomics Centre, Victoria, BC, Canada, V8Z 7X8; Division of Medical Sciences, University of Victoria, Victoria, BC, Canada, V8W 2Y2; Department of Biochemistry and Microbiology, University of Victoria, Victoria, BC, Canada, V8W 2Y2; Department of Medical Oncology, BC Cancer – Victoria, Victoria, BC, Canada, V8R 6V5; Department of Medicine, University of British Columbia, Vancouver, BC, Canada, V6T 1Z3

**Keywords:** Intermittent fasting, time-restricted eating, cancer, autophagy, metabolites

## Abstract

Emerging preclinical data suggests dietary interventions, including intermittent fasting, may play a key role in altering cancer progression. In a feasibility trial monitoring cellular, clinical, and qualitative changes, ten patients with chronic lymphocytic leukemia followed time-restricted eating for three months, and five patients for six months. Seven of fifteen participants (47%) experienced a decrease or stabilization in malignant lymphocyte counts on time-restricted eating, while malignant lymphocyte accumulation slowed in five participants (33%) or had no effect in three participants (20%). A reduction in malignant lymphocyte counts were accompanied by an unexpected decline in cellular autophagy in malignant lymphocytes. Metabolite profiling identified microbial-derived bile acid metabolites, glycoursodeoxycholic, taurochenodeoxycholic, glycolithocholic and ursodeoxycholic acid, and short-chain fatty acids, dehydrolithocholic and apocholic acid, that changed with time-restricted eating. Participant quality of life improved during time-restricted eating. These results connect time-restricted eating with shifts in autophagy, microbial-derived metabolites, tumor progression, and quality of life.

## INTRODUCTION

Systems-wide metabolic derangements, at the cell, tissue, and organismal levels, are a central feature of cancer progression (Hanahan & Weinberg, 2000; Faubert et al., 2020). To tackle perturbations at all three of these levels, dietary modifications have increasingly become a promising avenue for abating cancer proliferation (Antunes et al., 2018; Steck & Murphy, 2019). The pretext of using diet as a cancer treatment strategy is that sensitizing cancerous cells to treatment may occur through manipulating the energy metabolism (Madkour et al., 2020; Mindikoglu et al., 2020), the tumor microenvironment (TME), epigenetics, and the microbiome (Shi et al., 2021; Ahmad et al., 2023).

Intermittent fasting (IF) is a diet-based therapy that alternates between periods of free eating and abstaining from food and, sometimes, drinks (Clifton et al., 2021; Longo & Mattson, 2014). IF is of particular interest because it stimulates evolutionary conserved, adaptive cellular responses that are integrated between and within organs in a manner that improves glucose regulation, increases stress resistance, and suppresses inflammation (de Cabo & Mattson, 2019; Patterson & Sears, 2017). In particular, the cyclical oscillation of nutrient availability in time-restricted eating (TRE), where intake is limited to a defined number of daytime hours, takes advantage of the highly orchestrated systemic and cellular responses of the metabolic switch (de Cabo & Mattson, 2019; Di Francesco et al., 2018).

One of the cellular responses to nutrient supply is autophagy, which plays pleiotropic roles in cellular and organismal homeostasis (Galluzzi et al., 2015). Basal autophagy is crucial for cellular and tissue homeostasis to mitigate physiological stress through the removal of damaged organelles, protein quality control, and genomic integrity. Autophagy activation serves as a survival mechanism against antitumor treatments (Chen & Karantza, 2011) as well as during times of nutrient restriction. In the latter scenario, autophagy breaks down intracellular reserves of macromolecules, such as proteins and lipids, into amino acids and fatty acids that, together, can provide precursors for bioenergetic and biosynthetic needs (Lum et al., 2005). However, upregulation of autophagy can promote cell death, and the balance with survival appears to be context dependent. Nevertheless, inhibiting the pro-survival functions of autophagy remains an attractive anti-cancer strategy (Steck & Murphy, 2019; Chen & Karantza, 2011; Amaravadi et al., 2016). Nutritional interventions, including fasting mimetics (Nencioni et al., 2018), time-or caloric-restricted diets, and their impacts on autophagy and cancer, are an active area of study (Stringer et al., 2022).

The microbiome communities of the gut and resulting gut-associated metabolites are emerging as key contributors in host immune factors, neoplasia, and response to therapy (Rajpoot et al., 2018; Lynch & Pedersen, 2016). Influenced by both endogenous and exogenous factors, alterations in the composition and function of these intestinal microbes are demonstrated to play a major role in the pathogenesis of various neoplasms (Rajpoot et al., 2018; Gopalakrishnan et al., 2018) necessitating a paradigm shift to viewing microorganisms as an endogenous epidemiological exposure (Hamada et al., 2019). Animal models suggest that IF may be a modulator of microbial balance (Hu et al., 2023; Ridaura et al., 2013), results that are beginning to be mirrored in human trials. The restructuring of the microbiome may have direct consequences on metabolic dysbiosis of various compartments, including the plasma, gut, and cecum (Shi et al., 2021). To our knowledge, no studies have yet been completed that tests the impact of TRE on the gut microbiome in individuals with cancer, but research in other populations has found TRE to increase microbial richness (Su et al., 2021) and result in substantial remodeling of the gut microbiota (Ozkul et al., 2020).

Chronic lymphocytic leukemia (CLL) is a mature B cell neoplasm characterized by a progressive accumulation of monoclonal B lymphocytes. CLL and small lymphocytic lymphoma (SLL) are considered one disease with different manifestations. The malignant cells in CLL and SLL have identical pathologic and immunophenotypic features, with the difference being the location of involvement in the blood versus lymph nodes, respectively (Childs et al., 2019). CLL is the most common adult leukemia in Western countries, accounting for approximately 25 to 30 percent of all leukemias. CLL/SLL is most often diagnosed in older adults, with a median age at diagnosis of approximately 70 years. It is indolent by nature, resulting in a median survival of 12-15 years with current therapies (Aksungar et al., 2007). Most patients have an initial period of surveillance as they do not require active treatment. The clinical course is characterized by a slow progression of peripheral lymphocyte count and disease burden over time until the disease is impacting the quality of life (QOL), causing symptoms, or threatening organ function, at which time chemotherapy is recommended. The clinical characteristics of this patient group provide an ideal population for dietary interventions such as IF, without the confounding influence of anti-cancer treatment.

The goal of this study was to assess the feasibility of conducting a TRE study in patients with CLL and assess for a signal on tumor control, autophagy induction, and microbial-metabolite composition.

Secondary outcome measures include dietary adherence and acceptability, patient safety data including changes in body weight, side effect monitoring, weekly nutrition assessments, and QOL measures. To our knowledge, examination of the role of TRE-induced autophagy in patients and how this may impact tumor control is limited (Aksungar et al., 2007). Our results demonstrate the safety and acceptability of the diet while establishing a connection between dietary TRE with improvements in cancer control and alterations in autophagy, QOL, and microbial-derived metabolite levels.

## RESULTS

### Participant demographics

All participants met the inclusion/exclusion criteria and completed informed consent. A recruitment rate of 77% was achieved: 22 patients were invited to participate, five declined, two were removed due to starting chemotherapy, and 15 completed the entirety of the study (Figure 1). Participant characteristics are described in Table 1. In summary, at the time of recruitment all participants were of Rai (Stauder et al., 2017) stage 0 disease, median age was 69 years, 40% were female, and 60% male.

**Figure 1.**
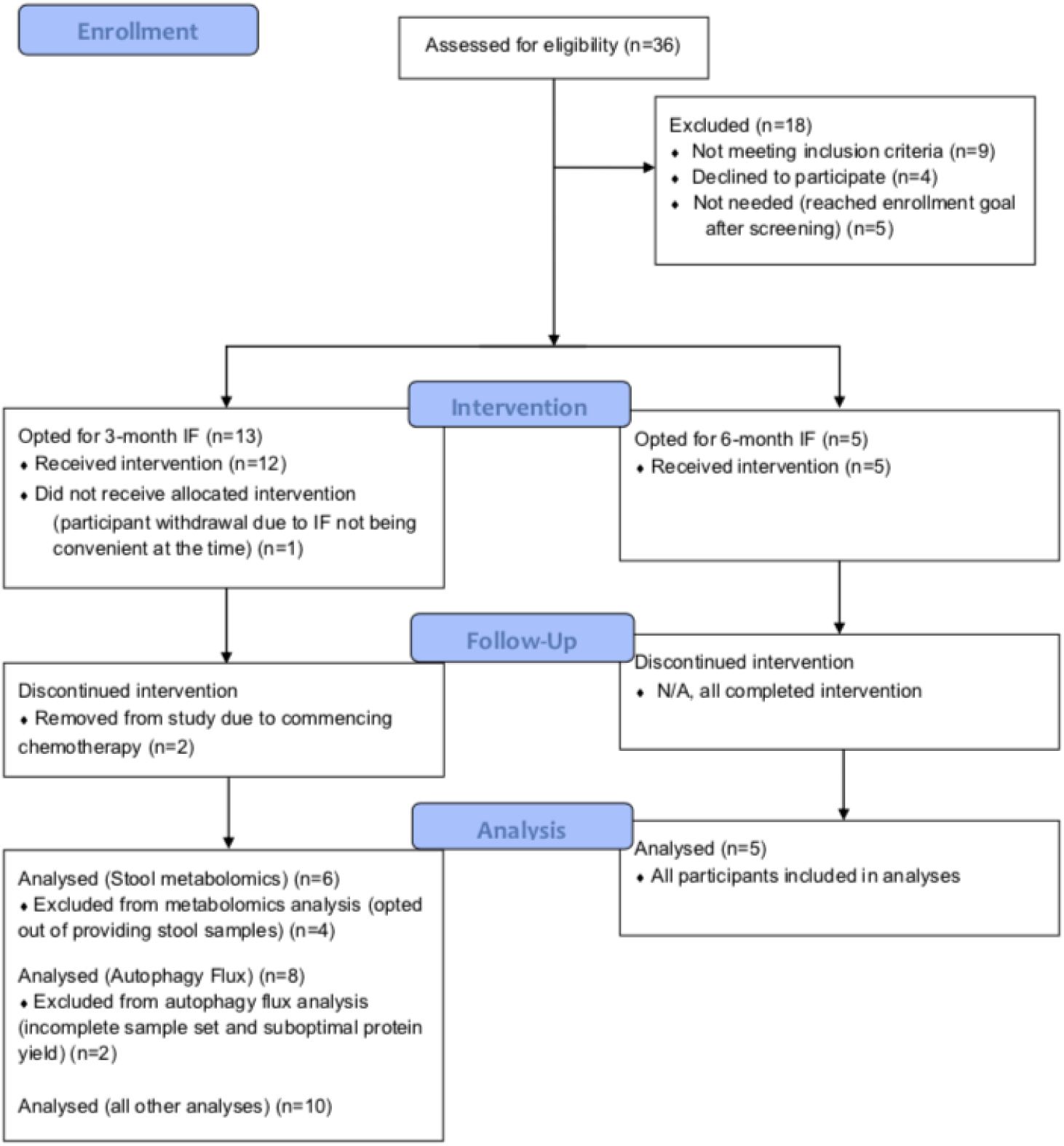
CONSORT diagram demonstrating the flow of participants through the study, including the three and six-month time-restricted eating parallel interventions.

**Table 1.**
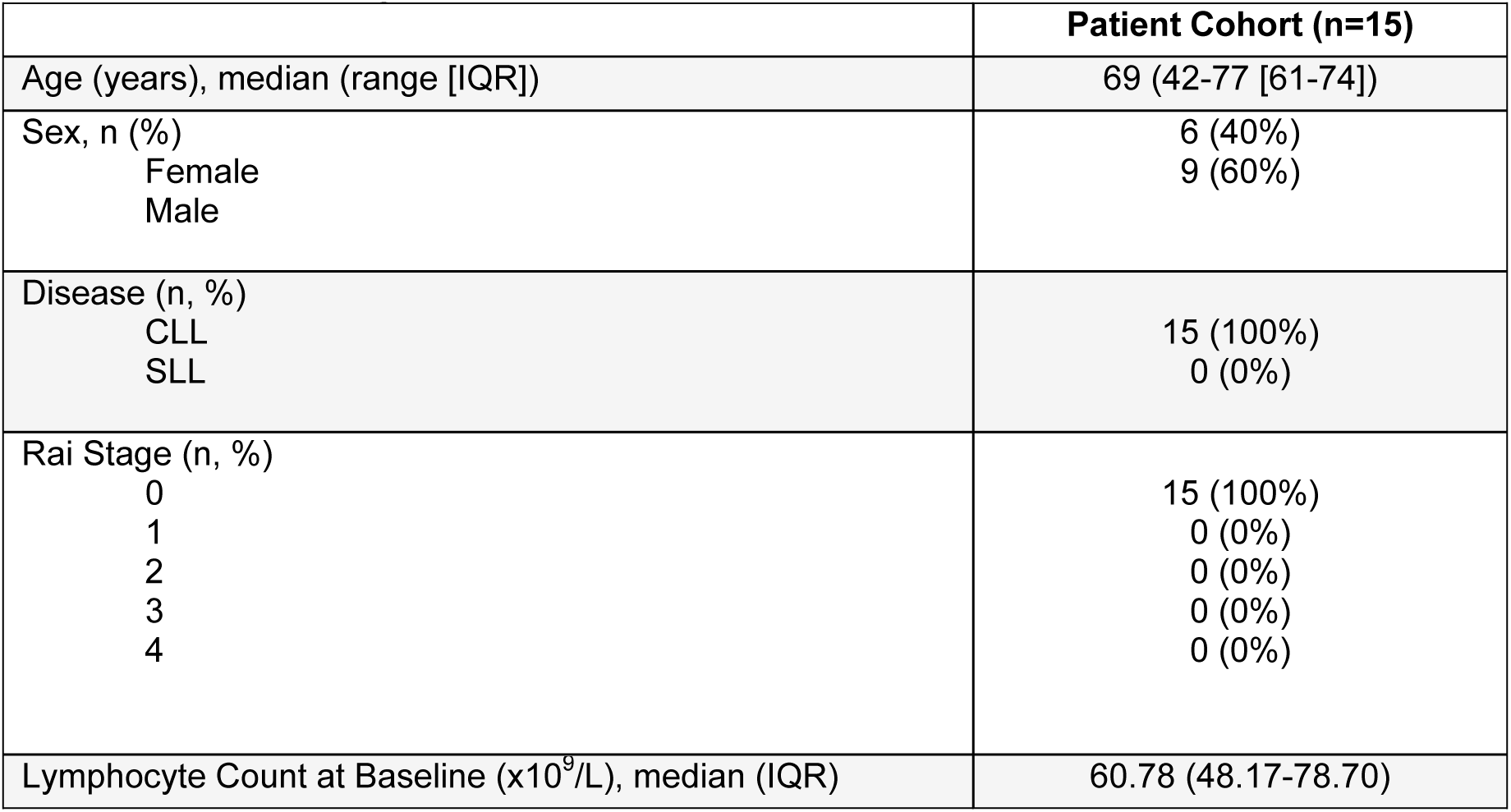
Patient demographics at time of recruitment.

### Time-restricted eating is acceptable and safe

Ten participants completed three months (90 days) of fasting, while five completed six months (180 days). Overall adherence was 99.2%, with a single participant taking two extra days off the intervention, beyond what was permitted in the protocol. The average eating window was 7 hours 28 minutes (median 7 hrs 45 mins, range 6 hrs 10 mins - 8 hrs) and all participants delayed their usual initiation of food intake but completed their eating by 8:00 PM (Table 2), per study protocol. Forty percent of participants, all from the three-month fasting cohort, did not take any days off the TRE intervention, voluntarily adhering to a feeding window of less than eight hours per day for the entirety of the study. All participants who fasted for six months took at least two days off TRE intervention for two weeks, which was permissible per study protocol.

**Table 2.** Dietary adherence rates.

| Participant ID | Fasting duration (months) | Mean daily fasting duration (hr:min) | Shortest (hr:min) | Longest (hr:min) | # 1 offs /wk (%) | # 2 offs /wk (%) | Total off wks (%) |
| --- | --- | --- | --- | --- | --- | --- | --- |
| S01 | 3 | 7:48 | 7:32 | 7:55 | 0 (0) | 0 (0) | 0 (0) |
| S02 | 3 | 7:15 | 6:30 | 8:00 | 0(0) | 0 (0) | 0 (0) |
| S03 | 3 | 8:00 | 8:00 | 8:00 | 4 (31) | 4 (31) | 8 (62) |
| S04 | 3 | 6:10 | 5:30 | 7:30 | 0(0) | 0 (0) | 0 (0) |
| S05 | 3 | 8:00 | 7:30 | 8:00 | 4 (31) | 1 (8) | 5 (38) |
| S06 | 3 | 7:28 | 6:15 | 8:00 | 0(0) | 0 (0) | 0 (0) |
| S07 | 3 | 7:45 | 7:20 | 8:00 | 9 (69) | 1 (8) | 10 (77) |
| S08 | 3 | 8:00 | 7:15 | 8:00 | 0 (0) | 1 (8) | 1 (8) |
| S09 | 3 | 8:00 | 6:30 | 8:00 | 0 (0) | 0 (0) | 0 (0) |
| S10 | 6 | 7:41 | 6:00 | 8:00 | 2 (8) | 0 (0) | 2 (8) |
| S11 | 3 | 6:35 | 4:48 | 8:00 | 0 (0) | 0 (0) | 0 (0) |
| S12 | 6 | 6:49 | 4:30 | 7:30 | 8 (31) | 0 (0) | 8 (31) |
| S13 | 6 | 7:52 | 7:20 | 8:00 | 7 (27) | 2 (8) | 9 (35) |
| S14 | 6 | 8:00 | 7:30 | 8:00 | 8 (31) | 2 (8) | 10 (39) |
| S15 | 6 | 7:46 | 6:16 | 8:00 | 4 (15) | 0 (0) | 4 (15) |
| TOTAL |  |  |  |  |  |  |  |
| Mean |  | 7:32 | 6:35 | 7:55 | 3.07 | 0.73 | 3.8 |
| Median |  | 7:46 | 6:30 | 8:00 | 2 | 0 | 2 |

During the weekly dietitian check-ins, participants provided qualitative feedback on their ability to comply with and maintain the diet. No participants expressed dissatisfaction or struggled to maintain the eating pattern. Several participants noted that a few weeks were required before the TRE felt routine, but no one required additional reminders or encouragement from the study team to adhere. Participants who took the most days off intervention followed TRE during the summer months, expressing the challenge of discontinuing intake by 8:00 PM during this season due to the extra socializing later in the evenings (e.g., vacation). When invited to participate, all participants scheduled their start date to avoid overlapping with December holidays.

Participant safety, as pre-defined by Expanded View Table 3, was assessed on a weekly basis for the first six weeks of the study and then every other week thereafter. Participants were graded for each domain of the safety assessment, designed to align with The European Organisation for Research and Treatment of Cancer (EORTC) Quality of Life Questionaiire-C30 version 3 (QLQ-C30) (Aaronson et al., 1993) domains - physical tolerance (including weight change), thoughts and feelings, and concerns - then assigned an overall score. Seven participants (47%) scored “good/great” during the entire study, while an equal number of participants (47%) received a score of “satisfactory” for one week of the study but scored “good/great” for the remainder of the study. One participant scored “satisfactory” for four recurring weeks due to weight loss, but the remaining eight weeks scored “good/great.” Of the satisfactory scores, four were judged to be less than “good/great” due to weight loss (1.5-2% weight loss over the preceding week), two were due to other coinciding health issues (e.g., acute COVID-19 infection), and two were due to transient light headedness. Two participants were removed early in the study due to CLL progression requiring treatment. One participant experienced a cardiac event despite having no pre-diagnosed cardiac history, otherwise no adverse events were reported. One week of TRE was associated with a mean weight loss of 0.39 lbs, though this correlated with participants weight goals, from +5 lbs to -20 lbs (+3.1 to -10.5% change in total body weight over three months) and -3 lbs to 14.5 lb (-1.7 to -8.1% change in total body weight over six months) (Table 3).

**Table 3.** Changes in body weight over TRE period.

| Participant ID | Fasting duration | Start weight (lbs) | End weight (lbs) | Absolute difference (lbs) | Total % lost | % lost/month |
| --- | --- | --- | --- | --- | --- | --- |
| S01 | 3 | 172.2 | 168.8 | 3.4 | 2.0 | 0.66 |
| S02 | 3 | 208 | 206 | 2 | 1.0 | 0.32 |
| S03 | 3 | 192 | 190.8 | 1.2 | 0.6 | 0.21 |
| S04 | 3 | 196 | 185 | 11 | 5.6 | 1.9 |
| S05 | 3 | Incomplete data |  |  |  |  |
| S06 | 3 | 190 | 170 | 20 | 10.5 | 3.5 |
| S07 | 3 | 160 | 165 | -5 | -3.1 | -1.0 |
| S08 | 3 | 224 | 222 | 2 | 0.9 | 0.3 |
| S09 | 3 | 266.6 | 262.9 | 3.7 | 1.4 | 0.5 |
| S10 | 6 | 178 | 163.5 | 14.5 | 8.1 | 1.4 |
| S11 | 3 | 220.9 | 211 | 9.9 | 4.5 | 1.5 |
| S12 | 6 | 146 | 141 | 5 | 3.4 | 0.6 |
| S13 | 6 | 172.5 | 169.5 | 3 | 1.7 | 0.3 |
| S14 | 6 | 224 | 215 | 9 | 4.0 | 0.7 |
| S15 | 6 | 161.8 | 152.9 | 8.9 | 5.5 | 0.9 |

### Longer periods of time-restricted eating are associated with a greater reduction in lymphocyte counts

For each of the 15 participants, a scatterplot demonstrated changes in absolute MLC over time, with lymphocyte count on the *y*-axis and time (in months) on the *x*-axis (Figure 2A). The data are categorized into three treatment phases: pre-intervention, intermittent fasting, and follow-up. The blue dots represent the MLC prior to the start of TRE, the gold dots represent MLCs during TRE, and green dots represent MLCs post-TRE when patients had returned to their regular, *ad libitum* eating patterns. There were three groups of participants: in the left panel, participant followed the three-month TRE and were observed post-TRE with monthly blood draws for three months; in the center panel, participants followed a six-month TRE with no post-TRE follow-up; and in the right panel, participants followed three-month TRE and were observed for one month after completing the TRE. Both groups in the three-month TRE had the same intervention, varying only in their post-TRE observation length. Initially, it was theorized that a one-month follow-up period would be sufficient to understand the lingering effects of TRE; however, as data came in, it became evident that a longer follow-up period would be ideal. Consequently, the study protocol was amended. Visually observing the MLCs in these 15 participants (Figure 2A) from pre-intervention through TRE, most do not appear to have a significant change in the trajectory of their MLCs. However, participants S10, S12, S13, and S15 show a slower accumulation of their MLCs.

**Figure 2.**
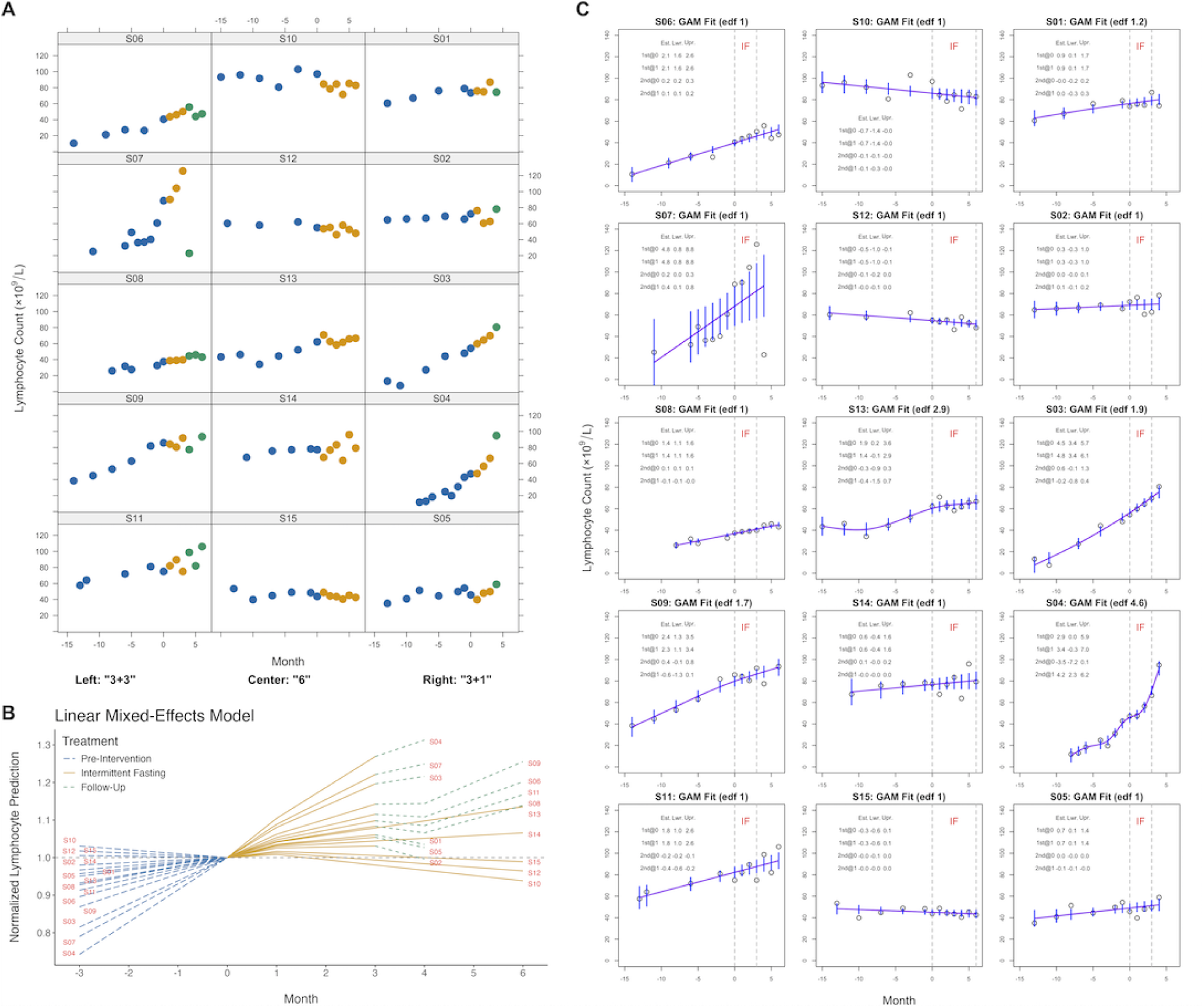
Plots illustrating the relationship between malignant lymphocyte count and time-restricted eating over time. **(A)** Scatterplot of absolute malignant lymphocyte count (MLC) over time with TRE. Blue dots represent MLC pre-intervention, gold dots during the TRE intervention, and green dots post-TRE. Left panel: three-month TRE plus three-months post-TRE; middle panel: six-months TRE; right panel: three-month TRE plus one-month post-TRE. **(B)** Linear mixed-effects model of MLCs over time with TRE. **(C)** Generalized additive models (GAMs) shown with purple trajectories and 95% confidence intervals in blue. Time-restricted eating (TRE) periods are marked by vertical dashed gray lines. Tables in each plot present first and second derivatives’ estimates for Months 0 and 1, with their 95% confidence intervals. Note: "edf" refers to effective degrees of freedom.

These MLC data was then fit to the linear mixed-effects (LME) model (Figure 2B) and the generalized additive model (GAM) (Figure 2C) to examine the relationship between MLC and time with a curve on the scatter plot.

The LME model fits straight lines for each treatment phase, with unique intercepts and slopes for each participant. The intercept estimates the baseline lymphocyte count at Month 0, while the slope represents the rate of change in lymphocyte count. A positive slope indicates an increase in lymphocyte count over time, and a higher absolute value of the slope suggests a faster rate of change. Conversely, a negative slope indicates a decrease in lymphocyte count over time. The LME model (Figure 2B) illustrated that the MLC decreased for three (20%) participants (S10, S12, and S15), and stabilized in four (27%) participants (S01, S02, S05, and S14). TRE slowed the rate of MLC accumulation in five (33%) participants (S06, S08, S09, S11, and S13), with no apparent effect in three (20%) participants (S03, S04, and S07). Upon completing the TRE, the MLC continued to decline in seven of the ten participants who had post-TRE data. Additionally, participants who fasted for six months showed a slower trend of MLC growth rate compared to those who fasted for three months (95% CI = [0.18, 7.49], *p* = .047, *t*_43_ = 2.05, Bayes factor 0.98). When pooling participant data during the TRE intervention, the interaction effect between the time and phase is estimated as negative but not statistically significant (95% CI = [-1.49, 1.39], *p* = .94, *t*_130_ = -0.07, Bayes factor 2.69), suggesting that mean MLCs did not increase at a higher rate upon adopting the TRE pattern. Overall, these statistics suggest that TRE may slow MLC proliferation in a subset of CLL patients.

The GAM is a nonparametric model that fits flexible and smooth curves, such as smoothing splines, to data points. This flexibility allows GAMs to capture complex patterns revealing intricate dynamics like periods of rapid increase followed by stabilization or other non-linear trends over time. In this context, the slope of the tangent line (i.e., the first derivative) to a non-straight curve varies across data points. A positive slope still indicates an increasing lymphocyte count at that point, while a negative slope indicates a decreasing count. The second derivative measures the rate at which the curve’s slope changes, indicating acceleration or deceleration.

Four participants, S03, S04, S09, and S13, exhibited non-linear patterns in their estimated GAM curves (Figure 2C). The higher the effective degrees of freedom (edf), the more pronounced the curvature of the GAM. A near-straight line is effectively fitted when *edf* = 1. Participant S09’s pattern suggests a slower rate of MLC growth, while S13’s indicates movement towards stability. Participant S04 also exhibits a non-linear pattern suggesting greater variability in MLC after the onset of TRE. The minimal to no effect observed in most patients between baseline and day 30 in the GAM exploratory analysis suggests that a fasting period longer than 30 days is required to observe an effect in MLCs.

### Time-restricted eating induces progressive reduction in tumor cell autophagy

In the three-month TRE cohort, there was a gradual decline in autophagy flux throughout all time points as determined by the relative net increase in LC3-II expression. For the six-month TRE cohort, there was a small increase in autophagy flux between the first and second time points; however, similar to the three-month TRE group, autophagy activity subsequently decreased below baseline levels (Figure 3A-C).

**Figure 3.**
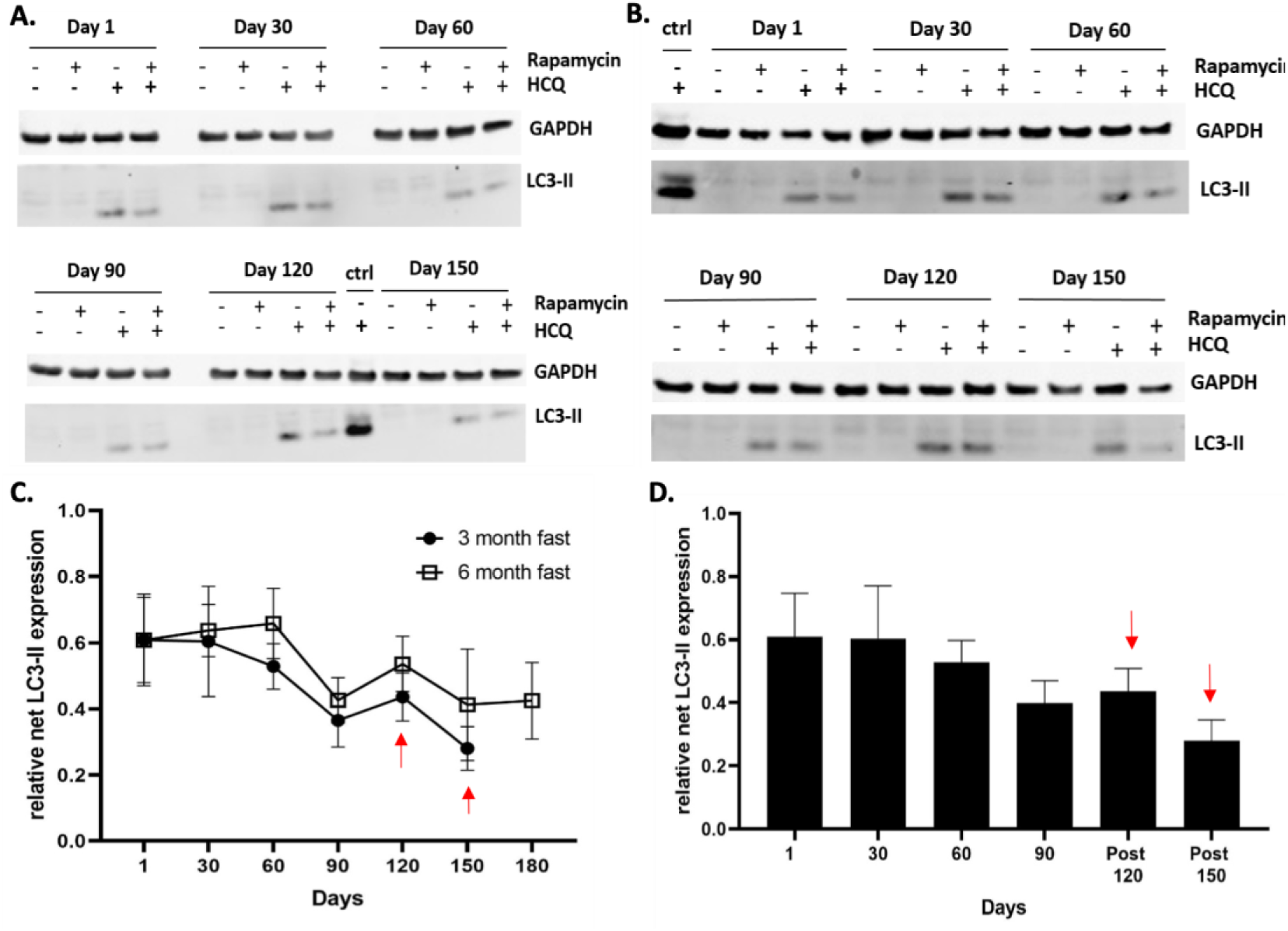
Autophagy flux analysis of the three-month and six-month intermittent fasting group show decreasing autophagy flux. Western blot analysis of **(A)** S11 (three-month) and **(B)** S12 (six-month). Cells were treated with or without hydroxychloroquine (HCQ, 30 μM) and rapamycin (0.40 μM). LC3-II was detected using an anti-LC3B antibody and GAPDH is shown as a loading control. Positive control lysates were obtained from healthy donor PBMCs treated with HCQ. **(C)** Time course of quantified LC3-II levels of all patients in the three-month TRE group versus six-month TRE group. Data points are expressed as the means +/- SEM of all patients in their respective cohort. Red arrows point to data points in the three-month cohort after TRE was completed. **(D)** Bar graph of three-month TRE cohort with follow-up monitoring after TRE was completed. Data points are expressed as the mean +/- SEM of all patients.

Participant S12, who had a pronounced decline in their MLC, also experienced the largest reduction in autophagy activity. Although the observed decrease was unexpected, autophagy activity continued to decline in three of the participants with longer-term follow-up data (Figure 3C, D). In addition, the relative LC3-II expression was higher at all time points in the six-month compared to the three-month TRE group (Figure 3C). In both groups, the sharpest decline in autophagy activity occurred between day 60 and 90. There was also a noticeable and higher level of total LC3-I and LC3-II conversion in control PBMCs from healthy donors compared to patients in the study. Thus, not only were the overall LC3 protein levels lower but there was also a general reduction in flux in all study participants.

To address whether there was an impact of TRE on autophagy activity for both the three-month and six-month TRE cohorts, the LME model was applied to the dataset. Despite observing changes within individual patients, an overall decreasing trend was observed when the data was examined in aggregate (95% CI = [-0.22, 0.04], *p* = .20, *t*_34_ = -1.31, and the Bayes factor 1.85 as anecdotal evidence supporting the null).

### Fecal metabolite abundances undergo shifts during three-month time-restricted eating

To examine whether TRE alters metabolite profiles, matched time course fecal samples collected from patients in both fasting groups were profiled for short-chain fatty acids (SCFAs; Expanded View Table 1) and bile acids (BAs; Expanded View Table 2), two of the major metabolite classes associated with microbial metabolism. In total, 82 targeted BAs and 11 SCFAs were profiled from each participant sample. In the three-month TRE cohort, three fecal samples were obtained: a baseline sample (day 1), an end-of-TRE sample (day 90), and a follow-up sample one month (day 120) following completion of the TRE (Figure 4A). An initial analysis of pre-TRE samples showed a diversity in metabolite composition across participants with no specific pattern or clustering (Figure 4B). A sub-analysis of the top 25 most significant SCFA and BA metabolites revealed several significant shifts before and after the TRE intervention. In one patient, S02, who experienced a clinically significant MLC decrease of 20.6% in one month, there was a dramatic reduction in the abundance of glycoursodeoxycholic acid between baseline and at the end of the three-month TRE time point, whereas the level of taurochenodeoxycholic acid levels increased from the baseline sample to the time of follow-up (Figure 4B). In two other participants who experienced progressive MLC declines, S09 and S11, the overall metabolite diversity remained stable over the TRE period.

**Figure 4.**
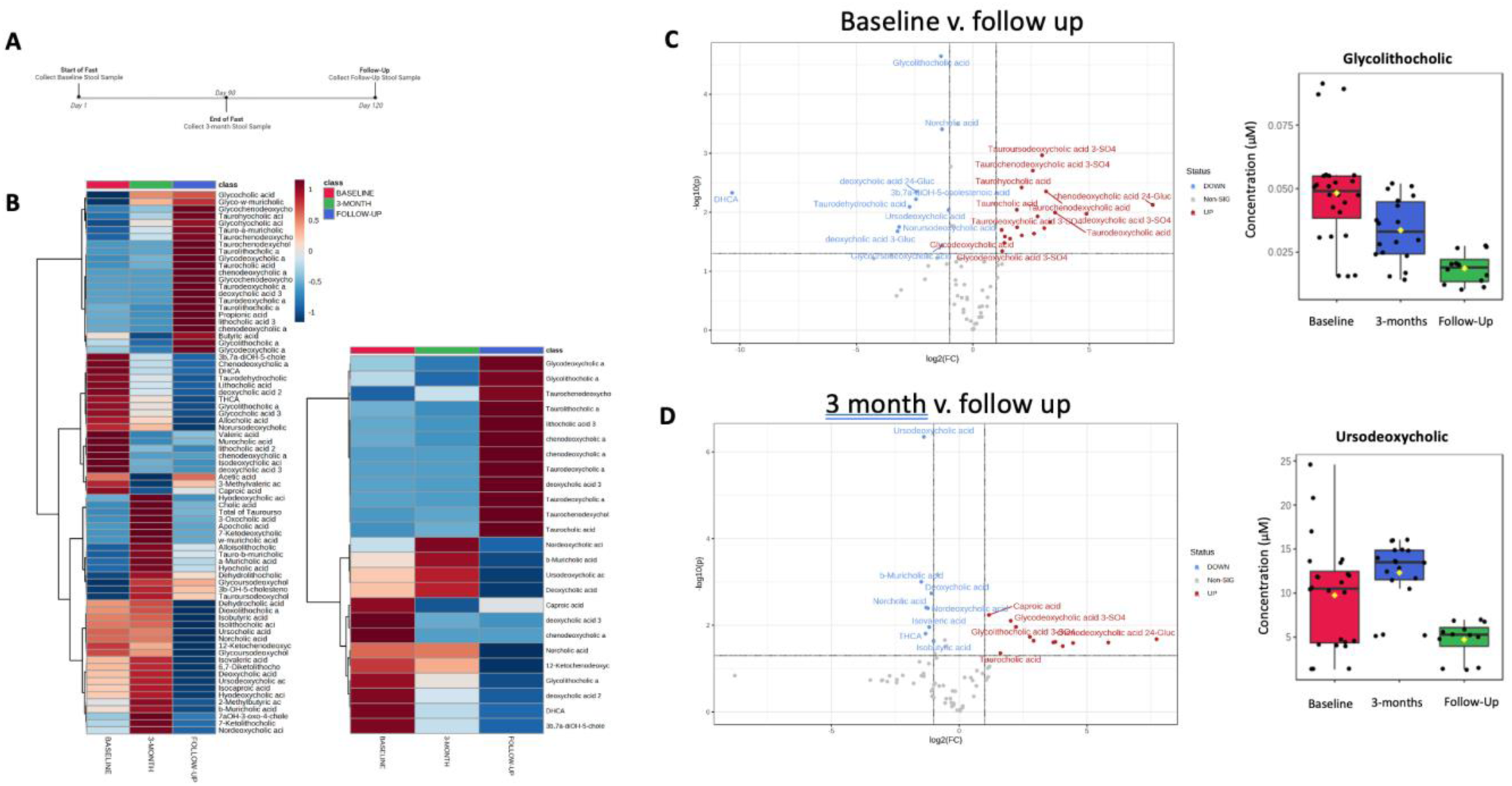
Metabolite profiling of three-month intermittent fasting group. **(A)** Timeline of stool collections for the three-month intermittent fasting group. **(B)** Heat map of metabolite abundance in the three-month TRE group. **Left:** Heat map of short-chain fatty acid (SCFA) and bile acid (BA) metabolites. Dendrograms representing Ward’s clustering of Euclidean distances among metabolites. Triplicate value calculated as the average of the duplicates. **Right:** The most significant 25 SCFA and BA metabolites ranked by t-test. Dendrograms representing Ward’s clustering of Euclidean distances among metabolites. Triplicate value calculated as the average of the duplicates. **(C)** Volcano plot of significant metabolites in blue (down)/red (up) (p<0.05*; fold change (FC) > 2), comparing baseline and follow-up time points. Box plots expressed as the mean +/- standard deviation of glycolithocholic acid (p<0.001***) across all time points. **(D)** Volcano plot of the significant metabolites in blue (down)/red (up) (p<0.05*, FC > 2), comparing three-month and follow-up time points. Box plots expressed as the mean +/- standard deviation of ursodeoxycholic acid (p<0.001***) across all time points.

Among all the metabolites examined across each participant, glycolithocholic acid decreased over the entire three-month TRE period, including the follow-up time one month after completion of TRE (Figure 4C). In contrast, some metabolites showed a biphasic change in abundance. Ursodeoxycholic acid levels became more enriched at the end of the three-month TRE but were lower at the time of follow-up, one month following completion of the TRE (Figure 4D). The decrease in glycolithocholic acid mirrored the general reduction in autophagy activity, whereas higher autophagy activity was associated with an increase in ursodeoxycholic acid.

### Fecal metabolite abundances undergo shifts during six-month time-restricted eating

Changes in concentration in the major classes of gut microbial metabolites were assessed in participants who adhered to six-months of TRE. Of the five participants who continued the TRE for six months, fecal samples were obtained at three timepoints: a baseline sample (day 1), mid-point (day 120), and end of fast (day 180) (Figure 5A). In three of these patients, S10, S14, and S15, there was a major reduction in a cluster of the top 25 metabolites from the baseline sample to the end of the TRE (Figure 5B). In particular, two SCFAs, dehydrolithocholic acid (Figure 5C) and apocholic acid (Figure 5D), were the most differentially enriched. Interestingly, patients S10, S13, and S15 in this six-month arm all showed a stabilization or reduction in MLC throughout the TRE intervention.

**Figure 5.**
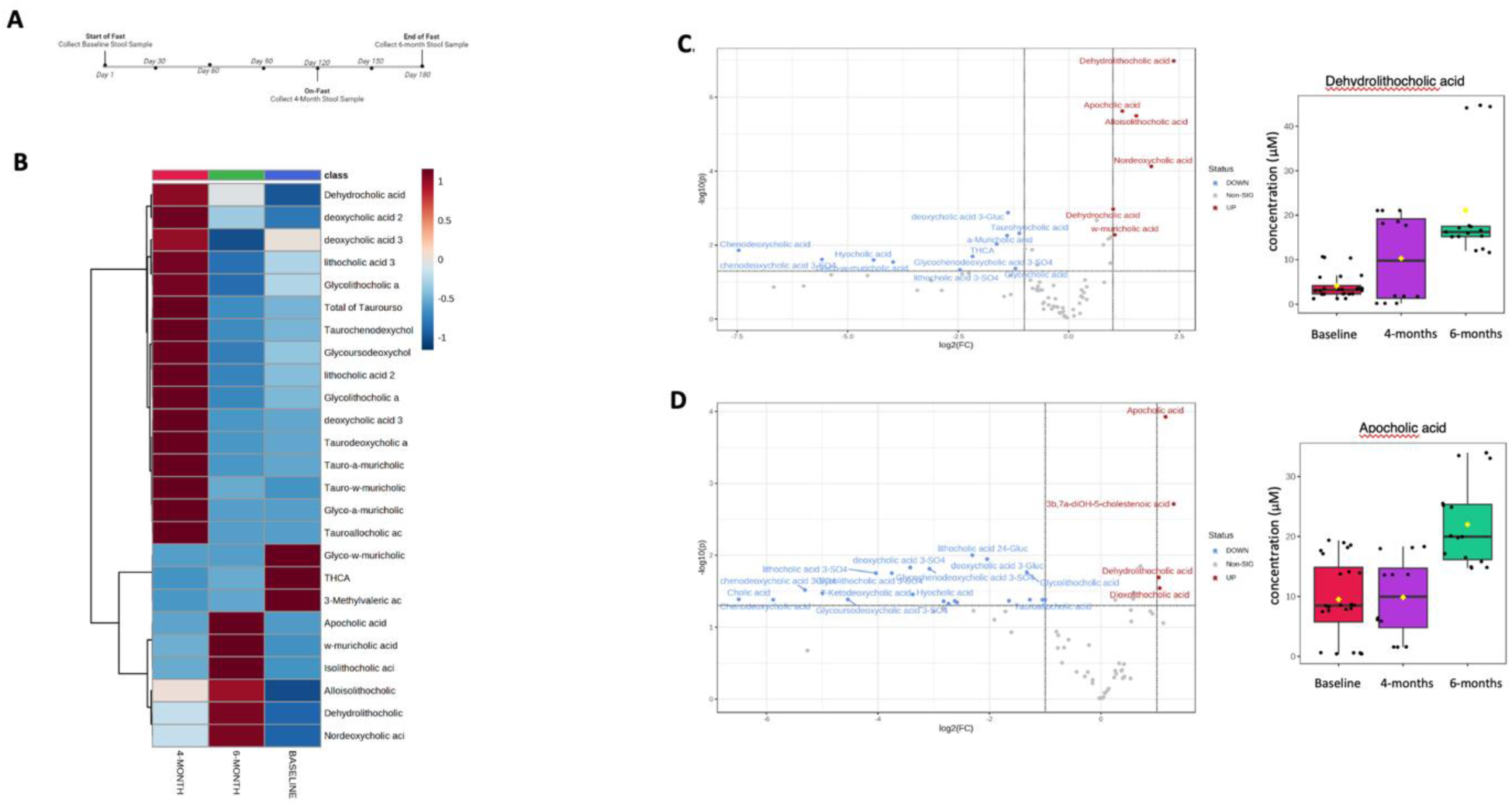
Metabolite profiling of the six-month intermittent fasting group. **(A)** Timeline of stool collections for the six-month cohort. **(B)** Heat map of metabolite abundance in the six-month cohort. The most significant 25 SCFA and BA metabolites calculated by ANOVA/t-test. Dendrograms representing Ward’s clustering of Euclidean distances among metabolites. Triplicate value calculated as the average of the duplicates. **(C)** Volcano plot of significant metabolites in blue (down)/red (up) (p<0.05*; fold change (FC) > 2), comparing baseline and six-months of fasting. Box plots expressed as the mean +/- standard deviation of dehydrolithocholic acid (p<0.001***) across all time points. **(D)** Volcano plot of the significant metabolites in blue (down)/red (up) (p<0.05*, FC > 2), comparing 4 months and 6 months of fasting. Box plots expressed as the mean +/- standard deviation of apocholic acid (p<0.001***) across all time points.

### Differential changes in fecal metabolites between three-months versus six-months time-restricted eating

Lastly, it was inquired whether specific metabolite changes could be detected between the two fasting groups which varied in length. Of all the metabolites measured through a targeted analysis, the levels of 3,4-dihydroxyhdyrocinnamic acid and 12-ketolithocholic acid showed a significant increase in overall abundance in the six-month TRE versus the three-month TRE cohort (Figure 6).

**Figure 6.**
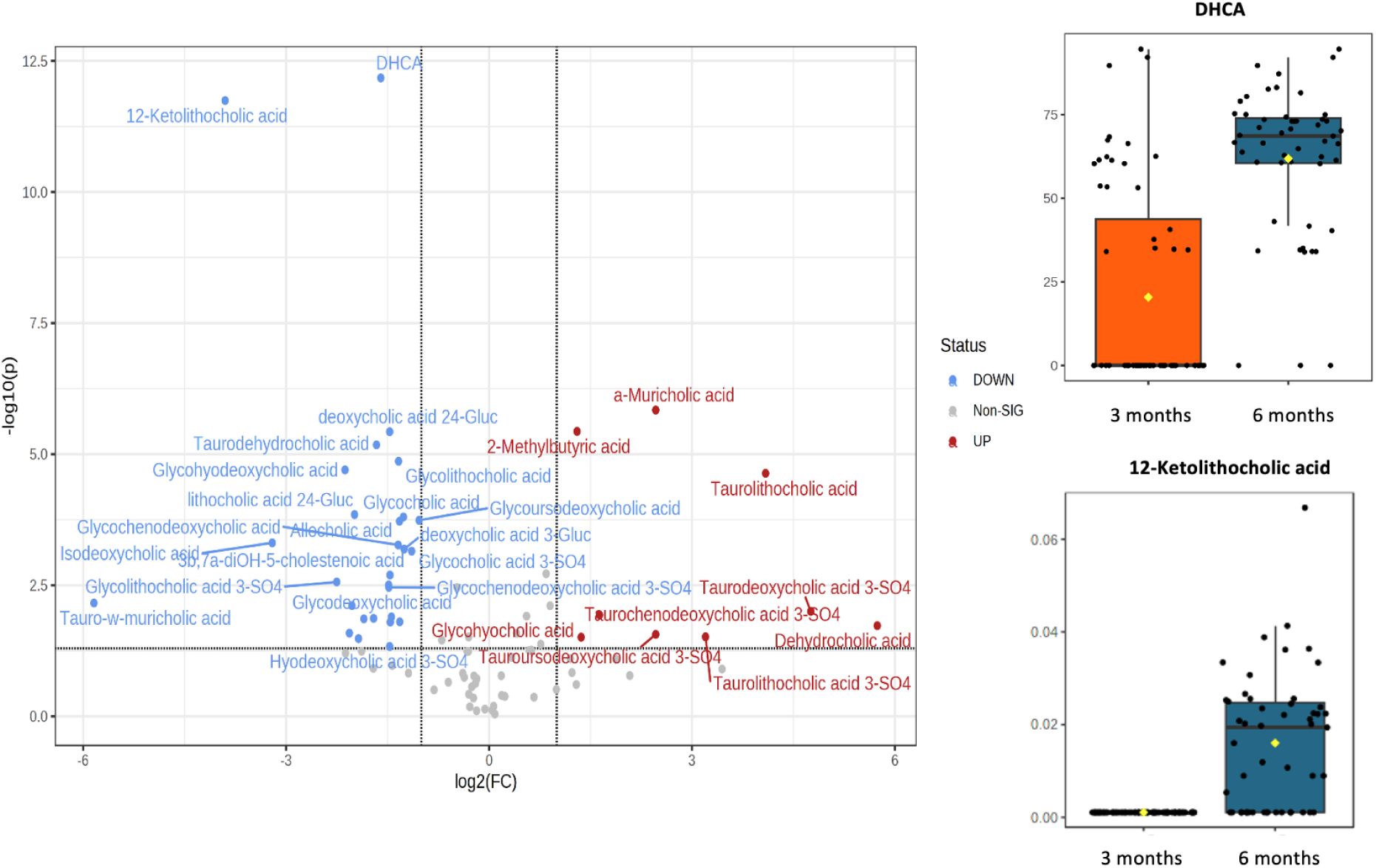
Metabolite profiling of three-month versus six-month intermittent fasting group. Volcano plot of significant metabolites in blue (down)/red (up) (p<0.05*; fold change (FC) > 2), comparing end of fast stool samples between the three-month and six-month intermittent fasting groups. Box plots expressed as the mean +/- standard deviation of 3,4-Dihydroxyhydrocinnamic acid and 12-ketolithocholic acid (p<0.001***) at three-month and six-month TRE.

### Time-restricted eating improves patient quality of life

All participants completed the QLQ-30 survey at baseline and at the end of the TRE intervention. Global health scores increased by a mean of 7.73 (95% CI = [1.54, 13.93]) points from 75.27 (95% CI = [66.67, 83.87]) to 83.00, *p* = .022. Of those (*n* = 8) who also completed the QLQ-30 one to three months after returning to their normal *ad libitum* diet, their global health score returned to baseline (95% CI = [-10.42, 10.17] for increment).

## DISCUSSION

At time of recruitment, all participants were medically assessed at Rai stage 0 CLL with no symptoms referable to active CLL and a stable weight history. Two participants were removed very early in the study, and one was removed during the follow-up, post-TRE period to begin treatment. While body weight measurements demonstrated a loss of 0.39 lbs per week, total percent weight loss ranged from +3.1% to -10.5%. Weight change reflected the weight goals of individual participants as well as their baseline weight. For example, individuals with overweight or obesity who desired weight loss represented the individuals who lost the greatest percentage of weight, while those within a healthy BMI maintained their weight, and one participant gained weight intentionally. This demonstrates that weight can be gained, maintained, or lost on TRE depending on one’s goals, food choices, and activity levels. Changes in body composition were not measured, but should be considered in future work, as maintaining lean body mass correlates with prognosis (Aduse-Poku et al., 2023; Au et al., 2021).

While the intervention did not provide any dietary guidelines, several participants used this as an opportunity to re-evaluate their food choices and align their eating with the AICR Cancer Prevention Recommendations (Collins, 2024). Several participants commented that engaging in this study introduced a new level of awareness to their eating habits and encouraged mindful and intuitive eating. This mindfulness may have assisted those with weight loss goals address any binge or emotional eating (Warren et al., 2017). During the weekly check-ins, patients reported an improvement in sleep, digestion, energy, and overall well-being; however, nearly all participants commented that they did not feel the QLQ-30 questions were sensitive to the changes they experienced because of TRE, but rather reflected personal and global affairs related to the COVID-19 pandemic. For example, question seven of the QLQ-30 asks whether the patient was limited in pursuing their “hobbies or other leisure time activities,” while question 22 asks whether they “worried.” Despite this limitation, several questions appeared sensitive to the changes because of TRE, such as those inquiring about sleep, fatigue, and overall health and QOL.

In this small pilot study, the EDA revealed seven participants (47%) who experienced a stabilization or reduction in MLCs during the TRE period which is of clinical interest. While the disease kinetics of CLL can follow various growth patterns, the most common being exponential unbounded growth and sigmoidal growth (Gruber et al., 2019), drops in MLCs are of particular interest. While the GAM only detected a potential reduction in MLC growth or disruptive effect in three participants, interventions that may disrupt cancer growth through a simple dietary manipulation alone are worth further investigation. Additionally, it is important to note that participants were not randomized based on disease severity or clinical status to undergo the three or six months of TRE. Given these limitations, further investigation in larger cohorts is warranted to validate these initial findings.

Given the field consensus that nutrient restriction activates survival programs that are mediated by autophagy (Lum et al., 2005), it was surprising that in some participants, autophagy activity declined over the time course of the TRE and was sustained upon return to *ad libitum* eating in three participants. One speculation is that TRE stimulates a subset of tumors to undergo autophagic tumor cell death, which initially may result in lower MLCs. Over time, cells may adapt to TRE and become insensitive to autophagy activation. One benefit of TRE in this scenario is that malignant lymphocytes may no longer have sufficient capacity to activate protective autophagy when exposed to systemic or molecular targeted therapy. Thus, TRE could synergize with other treatments, a prospect that will require further investigation. Although there was a trend towards higher autophagy levels in the six-versus three-month cohort, this may simply reflect the small size of the two cohorts.

Our exploratory analysis showed no consistent metabolite clusters within the total patient group; however, some metabolite changes were apparent and have described tumor cell modulatory and immunoregulatory roles. Glycoursodeoxycholic acid is involved in bile acid metabolism and has been implicated in cardiometabolic diseases. In one study, the administration of glycoursodeoxycholic acid ameliorated western-diet-induced atherosclerosis (Huang et al., 2021) and also type 2 diabetes (Chen et al., 2023) by altering bile acids and the levels of gut microbiota. Its role in the persistence of tumor cells is not known, although high levels of this tertiary bile acid are associated with an increased risk of colon and colorectal cancer (Cheng et al., 2023; Cai et al., 2022). Glycoursodeoxycholic acid may also change the anti-inflammatory features in the TME by suppressing TNF-α production and impairing natural killer T cell recruitment (Cheng et al., 2023). The reduction in MLCs may also be associated with the observed increase in taurochenodeoxycholic acid which has been reported to possess anti-proliferative and apoptosis-inducing properties (Xhang et al., 2022). There is also evidence that taurochenodeoxycholic acid suppresses IL-1β in peritoneal macrophages but can also promote expansion of CD3+, CD4+, and CD19+ cells in the peripheral blood of mice (He et al., 2005; Hong, 2024). Some reports have suggested that higher levels of circulating glycolithocholic acid may be associated with increased incidence of colorectal cancer in women (Loftfield et al., 2022).

In patients in the six-month TRE cohort, no consistent changes in metabolite profiles were observed. However, analysis of all patients revealed two BAs, glycolithocholic acid and ursodeoxycholic acid, which decreased and increased, respectively, during the longer TRE. Much less is known about the biological action of glycolithocholic acid at the cellular level; however, ursodeoxycholic acid has been reported to have both antitumor and immune-modulatory functions (Shah et al., 2006; Yao et al., 2020). For instance, ursodeoxycholic acid synergizes with proteasome inhibitors to induce glioblastoma cell death (Yao et al., 2020) and has been shown to suppress colorectal tumor growth (Ahmad et al., 2022; Goossens & Bailly, 2019; Nguyen et al., 2018). Immunologically, ursodeoxycholic acid suppresses the secretion of anti-inflammatory cytokines, including TGF-β, TNF-α, IL-1, and IL-6 (51). Among the four metabolites that increased during the six-month fast (dehydrolithocholic acid and apocholic acid) or differentially increased between the three-month versus six-month TRE cohorts (dihydroxyhdyrocinnamic acid and 12-ketolithocholic acid), their roles in cancer and immunity have not been reported. Due to practical and clinical considerations, not all patients were able to provide fecal samples at all the time points, thus, limiting the conclusions that can be deduced from this work.

Although TRE studies have been reported in the literature, this study comparing three-month versus six-month TRE demonstrated safety, feasibility, and adherence in a subgroup of leukemia patients. In addition, the correlative analysis revealed an unexpected reduction in autophagic activity that is contrary to some preclinical studies and other TRE-related studies (Antunes et al., 2018; Shi et al., 2023; Pietrocola et al., 2016). While the precise reason for these differences is unclear, several of the human studies were conducted on healthy individuals without cancer, whereas the murine systems primarily involved xenograft models where the consequences of TRE may be highly impacted by the mutational landscape of the tumor. In future studies, it will be crucial to clarify the possible mechanisms of decline in lymphocyte counts or autophagy regulation due to TRE. These efforts may lead to better biomarkers to stratify patients who are the most likely to benefit from metabolic interventions such as TRE.

## Data Availability

The data used in the analyses are available via the Open Science Framework at https://osf.io/yv62c/. All R code is available via the Open Science Framework at https://osf.io/yv62c/. Complete plots can be viewed on the Open Science Framework website at https://osf.io/yv62c/. Any additional data will be made available from the lead contact upon request.

## ACKNOWLEDGEMENTS

Thank you to Heather Lockyer and Alejandra Raudales for assistance in data management. Thank you to the BC Cancer Foundation (F2005888 to E.S. and N.M.) and to the Canadian Institutes of Health Research (PJT192015 to J.J.L.) for funding this study.

## AUTHOR CONTRIBUTIONS

E.S. contributed to conceptualization, funding acquisition, data curation, investigation, methodology, project administration, and writing the original draft. J.J.L. contributed to conceptualization, funding acquisition, methodology, resources, supervision, validation, and writing (review and editing). N.M. contributed to conceptualization, funding acquisition, investigation, methodology, supervision, and writing (review and editing). Z.W. and F.N. led the data curation, formal analysis, validation, and visualizations.

Z.W. contributed to writing the original draft. F.N. contributed to reviewing and editing the writing. S.P. and N.C. contributed to formal analysis, investigation and writing of the original draft. J.H. and D.G. contributed to the investigation, methodology, resources, supervision, and reviewing and editing the writing. All authors agreed to submit the manuscript, read and approved the final draft and take full responsibility of its content, including the accuracy of the data and the fidelity of the trail to the registered protocol and its statistical analysis.

## THE PAPER EXPLAINED

***Problems*** CLL is prevalent, slow growing, and usually indolent disease which progresses until medical treatment is required. No lifestyle modifications are known to slow CLL progression.

***Results*** 15 patients with CLL followed time-restricted eating for either 3 or 6 months. Eighty percent of participants experienced a clinical response (i.e., slowed tumor progression), with a greater response in those who followed time-restricted eating for 6 months compared to 3 months. These changes were accompanied by a reduction of autophagy within the tumor cells, shifts in fecal metabolites, and improvements in quality of life.

***Impact*** This is the first study demonstrating the positive impact of time-restricted eating on tumor burden in patients with CLL while concomitantly investigating autophagy flux as a potential mediating mechanism. Additionally, it confirms the safety and feasibility of time-restricted eating in this group and describes changes in gut metabolites that may have immune modulating implications in cancer.

## EXPANDED VIEW

Expanded View Table 1: Metabolites Included in Short Chain Fatty Acid Panel

Expanded View Table 2: Metabolites Included in Bile Acid Panel

Expanded View Table 3. Clinical Safety Parameters

Expanded View Figure 1: Sample Collection Diagram

## METHOD DETAILS

### Study approval

Harmonized research ethics board approval was granted on May 26, 2020, with the University of British Columbia-BC Cancer Research Ethics Board as the harmonized board of record and the University of Victoria as the harmonized partner board (REB Number H19-03072). Written and dated informed consent was collected from every participant prior to study initiation. The study was prospectively registered with clinicaltrials.gov, ID Number NCT04626843.

### Intervention

All participants met the inclusion/exclusion criteria outlined in Table 4 Participants were recruited between February 16, 2021, and March 7, 2022. Participants met with a Registered Dietitian, E.S., who taught the principles of TRE, reviewed the study protocol, and answered questions. Participants were given the option of slowly reducing their eating window over a duration of nine days (or less) or immediately imitating an 8-hour eating window. Participants selected the timing of their eating window (i.e., early versus late morning start time) but were asked to conclude intake by 8 PM. Participants were permitted one day off intervention per week and two weeks with two days off intervention to grant flexibility, accommodate special events (e.g., weddings, birthday celebrations), and promote diet adherence.

**Table 4.** Participant inclusion and exclusion criteria.

| Inclusion | Exclusion |
| --- | --- |
| Diagnosis of CLL or SLL<br>Age $\leq 85$<br>Lymphocytes $\geq 40$ and $< 150$<br>Hemoglobin $\geq 100\text{g/L}$<br>Platelet $> 100 \times 10^9/\text{L}$<br>BMI of $\geq 20\text{kg/m}^2$<br>ECOG Performance Status $\leq 2$<br>Not on anti-lymphoma therapy within the past 3 months<br>Not receiving anti-lymphoma therapy and not expected to require initiation of anti-lymphoma therapy within the next 3 months | Patient unable to give consent<br>Patient on medications required to be taken with food during the fasting window<br>Pregnancy<br>Diabetes mellitus<br>BMI drop to $\leq 18.5\text{kg/m}^2$ at any time during study |

Participants recorded their first and last time of intake daily on a provided journal along with their Sunday weight. These journals were sent to the Registered Dietitian for weekly review of protocol compliance and weight changes. The dietitian contacted participants to assess tolerance and safety using a pre-defined checklist that aligns, categorically, with The European Organisation for Research and Treatment of Cancer (EORTC) Quality of Life Questionaiire-C30 version 3 (QLQ-C30) (Aaronson et al., 1993). Participants answers, comments, and feedback were recorded. Based on the answers, participants were assigned a traffic light score per the safety criteria (Expanded View Table 3). This check-in also served as an opportunity to answer participants’ questions and concerns and remind them of upcoming specimen collection due dates. Monthly bloodwork was reviewed by the dietitian, and concerns or abnormalities were escalated to the Medical Oncologist, N.M. N.M. spoke to participants who had medical concerns or questions and liaised with their community-based physicians when needed.

In the absence of standard-of-care bloodwork in the previous three months, blood samples were collected, prior to initiating the intervention, to confirm eligibility. All participants had bloodwork completed on day 1, 30, 60, 90, and 120 of the intervention. For participants who opted for extended follow-up or fast for six months, bloodwork was also taken on day 150 and 180. Stool samples were collected immediately prior to beginning the TRE (baseline), at end of the fast, and one month post-fast for those on the three-month TRE (Expanded View Figure 1, sample collection diagram).

### Sex as a biological variable

Both male and female participants were included in this study. Due to the small sample size of 15 participants, sex was not examined as a biological variable; however, this would be recommended for future studies with a larger sample size.

### Patient sample collection and processing

All specimens and clinical data were obtained with informed written consent under protocols approved by the University of British Columbia - BC Cancer Research Ethics Board and the University of Victoria (H19-03702). Blood samples were obtained through LifeLabs. Blood was collected in sodium heparin tubes for peripheral blood mononuclear cell (PBMC) processing and serum separation tubes for serum processing. PBMCs were processed using the Ficoll-Paque gradient method. Sterile PBS was added to the blood in a 1:1 ratio and layered onto sterile Cytiva™ Ficoll-Paque™ PLUS Media (Cytiva; cat. no. 17144003). The mixture was centrifuged at 2250 RPM for 10 minutes at room temperature. The PBMC layer was collected and spun in sterile PBS at 1400 RPM for 10 minutes at room temperature. ACK lysis buffer (ThermoFisher; cat. No. A1049201) was added to lyse the remaining red blood cells if the cell pellet appeared red. Cells were cryopreserved with CellBanker 2 (Amsbio; cat. no. 11914) and stored in liquid nitrogen. Serum separation tubes were spun at 2250 RPM for 10 minutes at room temperature. The supernatant (serum) was collected and stored at -80°C. Patients self-collected stool samples into an OMNIgene GUT (OM-200) and OMNImet GUT (ME-200) kit (DNA Genotek), per manufacturer instructions. Upon receipt, samples were aliquoted equally into cryovials for long-term storage at -80 °C.

### Autophagy flux assay

PBMCs were cultured with TexMACS (Miltenyi) medium supplemented with 3% heat-inactivated AB human serum (Sigma-Aldrich), 1% penicillin (10000 U/mL)/streptomycin (10000 μg/mL), and IL-2 (1e6 U/ml). To assess levels of autophagy, PBMCs collected from each time point were treated with and without the addition of 0.4 μM rapamycin (Cayman Chemical Company) and 30 μM hydroxychloroquine (Sigma). Cells were incubated for 20 hours at 37◦C and harvested for protein lysate described below.

Positive controls were obtained through healthy donor leukopaks (StemCell) treated with 30 μM hydroxychloroquine. A conventional flux assay was used to determine the change in LC3-I to LC3-II processing as a proxy for autophagy activity (Klionsky et al., 2021) Lysates were generated from bulk peripheral blood lymphocytes collected during the fasting period.

### Western blotting

PBMCs were lysed in RIPA buffer (50 mM Tris-HCl pH 8.0, 1% NP-40, 0.5% Na-deoxycholate, 50 mM NaCl, 0.1 % SDS) containing a complete protease inhibitor cocktail, and phosphatase inhibitor cocktail for 30 min at 4°C. After centrifugation at 13,000 g for 15 min at 4°C, supernatant was obtained and stored at −80°C. Thermo’s NuPage reducing agent (10X) and LDS buffer (4X) were added to the lysates. Samples were loaded in equal amounts onto 4%–12% gradient NuPage SDS-PAGE gels (Thermo). Protein was transferred onto nitrocellulose membrane and blocked (LiCOR) for an hour at room temperature.

Membranes were cut and incubated overnight at 4°C with either LC3B rabbit (Novus Biological) and GAPDH mouse (Novus Biological) antibodies. Membranes were washed four times for 5 minutes each time with PBS-T and incubated for one hour at room temperature with their respective secondary antibodies. Membranes were washed three times for 10 minutes each time with PBS-T and then stored in PBS. Membranes were imaged with LiCOR Odyssey. Densitometries were captured with ImageJ.

### LC-MRM/MS metabolite profiling on stool biospecimens

Stool samples were sent to the University of Victoria Genome BC Proteomics Centre for LC-MRM/MS metabolite profiling (Han et at., 2015) of SCFAs and BAs (see panel list in Expanded View Table 1 and 2).

#### Sample Preparation

Each sample was homogenized with the aid of two 4-mm metal balls at a shaking frequency of 30 Hz for 1 min three times on a MM 400 mixer mill, followed by sonication in an ice-water bath for 3 minutes. The samples were centrifuged at 4000 RPM for 20 minutes at 5 °C. The clear supernatants were used for the assays described below.

#### Bile Acids

A mixture of standard substances of all the targeted BAs was dissolved in an internal standard (IS) solution of 14 deuterated bile acids in 40% acetonitrile. This solution was serially diluted with the same IS solution to have 10-point calibration standard solutions. The concentration range of each bile acid was 0.000025 to 10 μM. Twenty microliters of the clear supernatant of each stool sample were diluted 20-fold with the IS solution. Ten μL aliquots of each standard solution and each resultant sample solution were then injected to run UPLC-(-) ESI-MS/MS with scheduled MRM, using the LC and MS operating parameters as described by Han et al. (2015). Concentrations of detected bile acids in the provided samples were calculated from the internal standard-calibrated, linear-regression calibration curves of individual bile acids with the analyte-to-internal standard peak area ratios within an appropriate concentration range for each bile acid.

#### SCFAs

A mixture of 10 C2 to C6 SCFAs was prepared in HPLC-grade ethanol. This solution was serially diluted to have 9-point calibration standard solutions in a concentration range of 0.0025 to 1000 μM for each SCFA. Twenty μL of each standard solution or the supernatant of each sample solution was mixed with 40 μL of 200-mM 3-nitrophenylhydrazine (3-NPH) solution and 40 μL of 150-mM EDC-6% pyridine solution. The mixtures were allowed to react at 40 °C for 35 min. After the reaction, an aliquot of 20 μL of each solution was diluted 10-fold with a pre-made internal standard solution containing ten 13C6-3-NPH labeled SCFAs. Five μL aliquots of the resultant solutions were injected to run UPLC-MRM/MS, according to the LC and MS parameters described by Han et al. (Han et al., 2015). Concentrations of SCFAs were calculated from the internal standard-calibrated, linear regression calibration curves of individual SCFAs with the analyte-to-internal standard peak area ratios within an appropriate concentration range for each SCFA.

Metabolomics statistical analysis was carried out using described GraphPad Prism 8. For the metabolomics data, duplicate profiling was conducted, and a third concentration value was calculated as the mean of the first two concentration data points for each metabolite. For statistical analyses of the metabolomic data, MetaboAnalyst 5.0 (Pang et al., 2021) was used to carry out ANOVA, multiple *t*-tests, and others as described within text.

### Statistics

Exploratory data analysis was conducted by the *xyplot* function from the “*lattice*” R package (Sarkar, 2008). Linear mixed-effects models were conducted by the *lme*, *lmer*, and *lmBF* functions from the “*nlme*”, “*lme4*”, and “*BayesFactor*” R packages, respectively (Bates et al., 2015; Pinheiro et al., 2023).

Generalized additive models were conducted by the *gam* and *derivatives* functions from the “*mgcv*” and “*gratia*” R packages, respectively (Wood, 2017; Simpson & Singmann, 2024).

